# Magnetoencephalography-based prediction of longitudinal symptom progression in Parkinson’s disease

**DOI:** 10.1101/2025.09.05.25335158

**Authors:** Josefine Waldthaler, Igori Comarovschii, Daniel Lundqvist

## Abstract

The progressive motor dysfunction of Parkinson’s disease (PD) has been linked to widespread functional changes within the basal ganglia-thalamic-cortical network, particularly in the beta frequency range (13-30 Hz). However, the longitudinal evolution of cortical neurophysiological changes and their relationship to clinical progression remain poorly understood, particularly when accounting for the aperiodic component of the neuronal signal.

We aimed to close this gap by conducting a longitudinal resting-state magnetoencephalography (MEG) study in 27 persons with PD and 30 healthy individuals with a mean follow-up time of four years. Clinical symptom progression was assessed using the annual change in MDS-UPDRS-III motor scale, adjusted for changes in dopamine replacement therapy. Neurophysiological changes were assessed using source reconstructed MEG data parcellated into 68 cortical regions, from which power spectra were parameterized to separate oscillatory peaks in the beta, alpha (8-12 Hz), and theta (4-8 Hz) frequency bands from the aperiodic component described by its exponent and offset.

Neurophysiologically, we observed that a steepening of the aperiodic slope in the left sensorimotor region was associated with a progression of rigidity, and that an increase in the aperiodic offset with an increase in bradykinesia. Peak beta power in parieto-temporo-occipital regions was elevated at baseline compared with healthy controls, correlating with less severe bradykinesia. This negative relationship weakened over time in patients with progressive bradykinesia but remained stable in non-progressors, suggesting an association with compensatory mechanisms.

Using partial least squares regression to predict future motor disease progression from baseline neurophysiological features, the model was able to explain 19.5 % of the variability in motor progression in an independent validation cohort consisting of 18 persons with PD, with the aperiodic features contributing most to the predictions.

Our findings demonstrate a close relationship between cortical PD-related neurophysiological alterations and longitudinal changes in symptom severity. The results emphasize the importance of separating aperiodic neural activity from periodic oscillations since we show that a progressive steepening of the aperiodic exponent in the sensorimotor region represented the most prominent PD-related longitudinal cortical change, potentially reflecting a progressive shift of the excitatory-inhibitory balance towards inhibition. Furthermore, our results challenge traditional interpretations of cortical beta oscillations as purely antikinetic and highlight the potential predictive value of simple resting-state neurophysiological features for predicting future disease progression in PD.

## Introduction

### Alterations of cortical oscillations in Parkinson’s Disease

Slowness of movement or *bradykinesia* and muscle stiffness or *rigidity* are core motor symptom of Parkinson’s disease (PD) traditionally considered a result of altered basal ganglia output to the motor cortex.^1,2^ One potential neuromarker of impaired motor functioning in PD is oscillatory brain activity in the beta frequency range (13-30 Hz). Increased beta band power at the level of the basal ganglia has been consistently linked to severity of bradykinesia and to treatment response in PD.^3–5^ These findings align well with the conceptualization of beta as an antikinetic rhythm governing the “status quo” over initiation of a new movement.^4^

The relationship between *cortical* beta oscillations and motor symptoms, on the other hand, is far less clear. Indeed, some findings contradict a straightforward interpretation of beta as a purely antikinetic rhythm. For instance, previous evidence suggests that cortical beta power in the central sensorimotor region may be highest in early, untreated PD while it may decrease later in the disease course, eventually falling below healthy levels with *lower* sensorimotor beta power scaling with *higher* motor symptom burden.^6,7^ Additionally, resting state cortical beta activity may *increase* or remain unchanged with successful amelioration of motor symptoms by dopamine-replacement therapy (DRT).^6,8–10^

Beyond the motor cortex, global oscillatory slowing across the neocortex^11^, and widespread changes in cortical oscillatory network organization have been described in PD.^12–15^ This generalized *slowing* results from power increases in low-frequency bands (e.g., delta (2 – 4 Hz), theta (5 – 8 Hz) and concurrent power reduction in higher frequency bands (e.g., alpha (8 – 12 Hz) and beta). Recently, slowing of cortical activity in PD has been shown to exhibit a spatial adverse-to-compensatory gradient from superior parietal regions to the inferior prefrontal cortex.^16^

### The aperiodic component of the neuronal signal in PD

Spontaneous neuronal activity consists of two physiologically distinct components: On the one hand side, periodic or oscillatory fluctuations and on the other side, aperiodic, i.e., non-oscillatory fluctuations. In the frequency domain, the aperiodic component can be described as the offset and slope of the power spectrum which tends to follow a 1/f power law distribution.^17^ Aperiodic activity is hypothesized to reflect neuronal spiking activity and the so-called excitatory-inhibitory (E/I) balance where a steeper slope can be interpreted as increased inhibition over excitation and vice versa.^18^ When controlling for this aperiodic activity using parameterization of the power spectrum, periodic activity (i.e., true significant rhythmicity or oscillations) can thus be identified as any spectral peaks rising above the aperiodic component and described with its amplitude (or power), peak frequency and width.^19^

Recent re-appraisal of the cortical activity in PD considering the aperiodic component, has led to the conclusion that the slope of aperiodic component of the neuronal activity spectrum is steeper in PD compared with healthy individuals across central sensorimotor, parietal, temporal and occipital regions, indicating a shift of the excitatory/inhibitory balance in the direction of increased inhibition.^20,21^ That we have previously shown and replicated in a second cohort that beta band activity in the sensorimotor cortex is not different from healthy individuals when controlling for this aperiodic component^10,22^ raises the question how much of earlier PD-related findings were due to the lack of parametrization of the power spectrum into periodic and aperiodic components (but see also ^16^).

### Aim of this study

Very few longitudinal MEG studies on PD exist and, to the best of our knowledge, all report on the same patient cohort without separating the periodic and aperiodic components of the neurophysiological signal.^7,14,23^ In this longitudinal MEG study, we therefore aimed to investigate changes in the oscillatory and aperiodic components of neuronal activity across the entire cortex in relation to clinical progression of PD over a period of four years controlling for general effects of ageing by inclusion of a matched healthy control group. Further, we aimed to build and externally validate a predictive model for future disease progression derived from simple resting-state neuronal activity measures.

## Materials and methods

### Participants

Participants were enrolled at the Swedish National Facility for MEG (NatMEG, https://natmeg.se/). The study protocol was approved by the responsible ethics committee (Swedish Ethical Review Authority, DNR: 2019-00542) and followed the Declaration of Helsinki. All participants gave written informed consent before participating. Please see Supplementary Material for in- and exclusion criteria.

At baseline, a total of 66 patients with PD according to MDS diagnostic criteria and 68 healthy age- and sex-matched individuals were included in the study.^24^ Three years after finishing the initial data collection, we offered a follow-up visit to participants. We decided to include two pre-selection criteria in the PD group based on available clinical follow-up data: Those who suffered from severe PD-dementia, were known to be wheelchair-bound or treated with advanced therapies (deep brain stimulation, intrajejunal levodopa pump) were primarily excluded. A flowchart for details about the recruitment process at follow-up can be found in **Supplementary Fig. 1**. Twenty-eight patients with PD (42% of the baseline cohort) and 32 healthy individuals (47%) were recruited for the follow-up study with a mean follow-up time of 4.1 ± 0.6 years (**Table 1**).

**Table 1:** Demographics and clinical characteristics of the healthy control group (HC) and Parkinson’s disease groups (PD)

| Characteristic | HC (N = 30)<sup>1</sup> | PD (N = 27)<sup>1</sup> | t-statistic<sup>2</sup> | P-value<sup>2</sup> |
| --- | --- | --- | --- | --- |
| Age (years) | 69.5 (8.7) | 66.1 (10.2) | t (55) = 1.34 | 0.2 |
| Sex | 15 (50%) | 12 (44%) | $\chi^2 = 0.02$ | 0.9 |
| Disease Duration (years) | NA (NA) | 9.1 (3.6) | — |  |
| Time to Follow-up (months) | 47.8 (4.6) | 49.6 (9.6) | t (55) = -0.92 | 0.4 |
| MoCA Baseline | 26.2 (1.9) | 27.3 (2.3) | t (55) = -2.01 | 0.052 |
| MoCA Follow-up | 26.5 (2.0) | 27.0 (2.3) | t (55) = -0.75 | 0.5 |
| MDS-UPDRS III Baseline | NA (NA) | 17.8 (9.5) | — |  |
| MDS-UPDRS III Follow-up | 1.1 (2.8) | 19.3 (9.7) | t (55) = -9.85 | <0.001 |
| LEDD Baseline (mg/day) | 0.0 (0.0) | 469.6 (270.2) | t (55) = -9.53 | <0.001 |
| LEDD Follow-up (mg/day) | 0.0 (0.0) | 857.0 (293.3) | t (55) = -16.02 | <0.001 |

<sup>1</sup>Mean (SD); n (%)
<sup>2</sup>Within-group paired t-tests: MoCA HC: t (29) = -1.09, p = 0.283; MoCA PD: t (26) = 0.82, p = 0.42; UPDRS PD: t (26) = -0.87, p = 0.391; LEDD PD: t (26) = -7.4, p <0.001
MoCA = Montreal Cognitive Assessment, MDS-UPDRS III = part III of the Movement Disorder Society-revised version of the Unified Parkinson's Disease Rating Scale, LEDD = levodopa equivalent daily dosage.

### Clinical assessment

Clinical assessments and MEG recordings were performed in on-medication state, i.e., after intake of regular dopamine replacement therapy and all participants confirmed that they felt “on” before starting the MEG recording. Motor function was assessed using the motor scale of the Movement Disorder Society-sponsored revised version of the Unified Parkinson’s Disease Rating Scale (MDS-UPDRS-III) immediately before or after recording the MEG data.^25^ The MDS-UPDRS-III score was further divided into its symptom-specific subscales, i.e., bradykinesia, rigidity, tremor, and axial symptoms (**Supplementary Material**). General cognitive ability was assessed with Montreal Cognitive Assessment (MoCA).^26^ Levodopa equivalent daily dosages (LEDD) were calculated according to ^27^.

To account for the confounding effect of medication adjustments on clinical progression, we developed LEDD-adjusted progression rates for the total MDS-UPDRS III and its subscores. Clinical progression was defined as the absolute change in MDS-UPDRS III scores from baseline to follow-up (Δ). Medication dose change was calculated as the relative change in levodopa equivalent daily dose. To estimate the relationship between medication changes and clinical outcomes, we empirically derived an alpha coefficient by fitting a linear regression model (**Supplementary Fig. 2**):

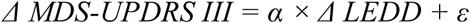

Using the estimated alpha coefficient, we calculated the annualized medication-adjusted motor progression as:

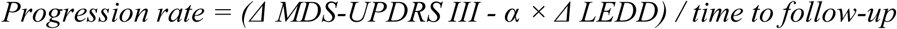

### Neuroimaging acquisition

The recording procedure was identical at baseline and follow-up and has been described in detail before.^22^ The MEG data was recorded on an Elekta Neuromag TRIUX 306-channel MEG system in a two-layer magnetically shielded room. The MEG system consists for 102 magnetometers and 204 planar gradiometers. Head position and any head movements were continuously recorded with head-position indicator coils (HPI), which were attached to the subjects’ heads and digitalized with Polhemus FASTRAK Motion tracker during the preparations. Electrooculogram (EOG) and electrocardiogram (ECG) were recorded to enable removal of muscle and heartbeat artifacts.

Eyes-open resting-state MEG data were collected using a at a sampling rate of 1000 Hz with an online 0.1 Hz high-pass filter and 330 Hz low-pass filter. Recordings lasted minimum 5 minutes and were conducted with participants in an upright position fixating a centrally presented crosshair. Data from one healthy participant had to be excluded due to technical failure during MEG recordings. Continuous head position was recorded during all except for two sessions (due to technical failure in one healthy control at baseline and one person with PD at follow-up). Empty-room recordings lasting at least 2 minutes were collected on the same day and were processed using the same pipeline to model environmental noise for source reconstruction.

Source spaces for MEG source reconstruction were created based on individual 3D T1-weighted magnetization-prepared rapid gradient-echo (MPRAGE) sequence structural MRI images (voxel size: 1x1x1 mm) obtained on a General Electric Discovery 3.0 T MR scanner at baseline, and a General Electric Signa-PremierXT scanner at follow-up, respectively. No structural MRI was available at baseline for seven participants due to restrictions during the Covid-19 pandemic (four HC, three PD), and at follow-up for seven participants with PD. Baseline images were used in these participants for source reconstructions at follow-up and vice versa. Note that at least one individual MRI scan was available for all participants except for two persons with PD in which an MRI template was used for source reconstruction.

### Neuroimaging data processing

Using MaxFilter software^28^, raw MEG data was pre-processed by applying temporal signal space separation with a buffer length of 10 s and a cut-off correlation coefficient of 0.95 with to suppress artefacts from outside the scanner helmet. Correction for head movements was done by shifting the head position to a position based on the median of the continuous head position.

Additional processing was executed with MNE-Python^29^ in Python 3.9. Data was 50 Hz-notch filtered to remove line noise, and band-pass filtered between 2 and 47 Hz. The continuous data were cut into 1.0 s epochs to subsequently reject epochs with extreme values or muscle artefacts using 5000 fT for magnetometers and 4000 fT/cm for gradiometers as cut-offs. An independent component analysis was performed using the *fastica* algorithm^30^ to identify and remove artefacts from blinks, eye movements (limited to a maximum of two components) and heartbeats (limited to a maximum of three components) based on their correlation with EOG and ECG data, respectively. This resulted in 3.31 ± 1.18 components to be rejected without evidence for a difference between groups or time points.

The individual T1-weighted MRI images were processed with Freesurfer (version 5.3) to obtain surface reconstructions of the cortical mantle using an automatic routine (*Freesurfer recon-all*). MEG data were co-registered to each individual’s segmented T1-weighted MRI using approximately 100 digitized head points. The cortex surface was parcellated into 68 ROIs (34 per hemisphere) according to the Desikan-Killiany Atlas.^31^ Noise-weighted minimum-norm estimates with dynamic statistical parametric mapping (dSPM)^32^ were used for source reconstruction with a source space consisting of 5,124 evenly spaced points sampled across the white matter surfaces. The inner skull boundary was estimated from the MRI to create a single shell volume conductor model. Time series from each ROI were extracted by singular value decomposition of all source points within the respective ROI.

### Parametrization of power spectra

The time series data were analysed for its the spectral properties by calculating the power spectral density (PSD) from 1 to 47 Hz across the entire ROI time series using Welch’s method by segmenting the continuous data into 5 s segments with 50% overlap. The aperiodic component of the power spectrum was estimated and subsequently separated from the periodic component (i.e., peaks rising above the aperiodic component representing putative oscillations) using the *fitting oscillations & one over f* (FOOOF) toolbox.^17^ This parameterizing algorithm fits a log-linear regression to the PSD which is subtracted before fitting Gaussian functions to the peaks in the PSD. The midpoint of the Gaussian function fitted to a given frequency band corresponds to the peak frequency in that frequency band and the height represents the signal power. The fitting parameters were set to a maximum of six peaks, peak threshold of 2 and a minimum peak height of 0.15, with a bandwidth between 1 and 10 Hz. The average R^2^ of the fit were 0.958 ± 0.028 at baseline and 0.960 ± 0.030 at follow-up for the PD group, and 0.947 ± 0.039 and 0.952 ± 0.032, respectively in the HC group. The estimated aperiodic component was subtracted from the full spectrum to obtain a flattened spectrum for each ROI. Spectral power peaks were initially identified for the theta (4-8 Hz), alpha (8-12 Hz), and beta frequency bands. However, given our relatively small sample and that oscillatory peaks are not expected in all frequency band across all cortical regions, there was a substantial amount of missing oscillatory peaks in the theta and alpha frequencies in many regions. In combination with the fact that the PD group exhibited a trend towards “alpha slowing” (i.e., shift of peak frequency from the lower alpha to the higher theta frequency range, **Figure 1**), we therefore decided to extend alpha to include the entire frequency range from 4-12 Hz. A version of the analysis considering alpha and theta frequency ranges separately can be found in the **Supplementary Material**.

**Figure 1:**
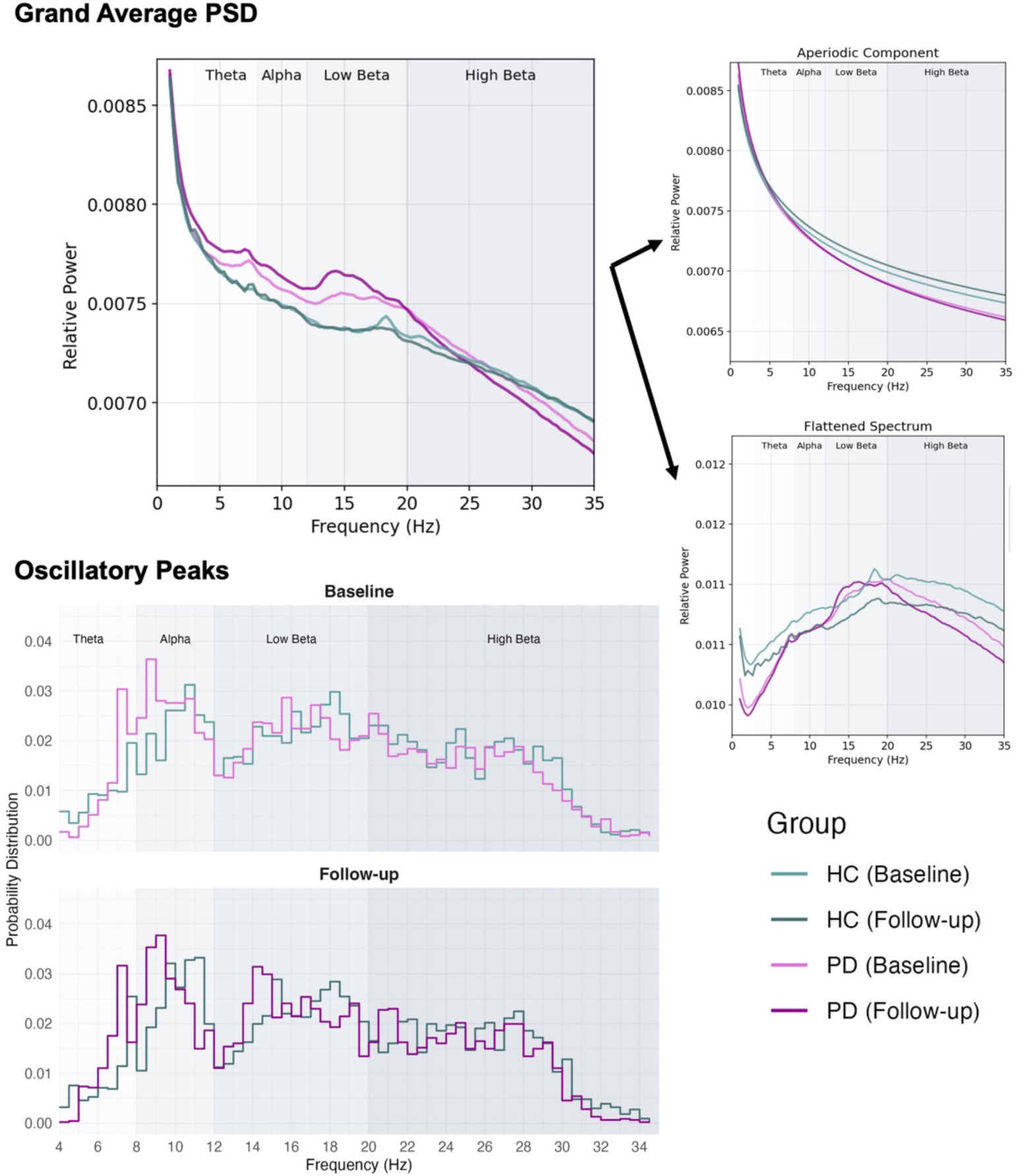
Left Panel: Grand average power spectrum across all regions by group and timepoint. The upper plot shows the full spectrum, the plots in the lower row show the aperiodic and periodic (“flattened spectrum”) components separately. **Right Panel: Histogram of the distribution of oscillatory peaks** across all regions by group and time point. Bin width 0.5 Hz. The beta band was extended to include frequencies up to 35 Hz for this visualization.

### Statistical Analysis

As described above, we computed the aperiodic component (exponent and offset) and peak frequency and power at peak frequency in two frequency bands (extended alpha and beta) from 68 ROI (in the following referred to as our variables of interest) from a total of 57 participants divided in two groups and at two timepoints.

Since no whole-cortex outcomes from this cohort at baseline have been reported before, we first compared baseline differences between the groups. To this end, we fit linear regression models with each outcome of interest as dependent variable and group as independent variable controlling for effects of age and sex:

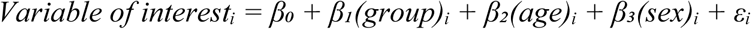

Likewise, associations of neurophysiological measures with motor performance at baseline in the PD group were inferred as follows:

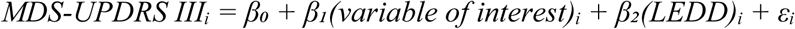

For group comparison of the longitudinal change in each of the pre-defined oscillatory and aperiodic variables of interest, linear mixed-effect models were fitted for each ROI using the lmer4 package in R^33^ with group*time interaction, age, and sex as fixed effects and subject as random effect:

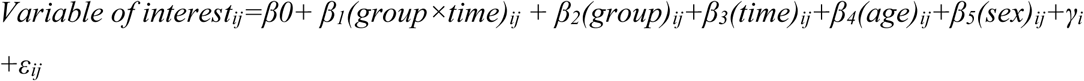

At baseline, time was set to zero, and at follow-up, time was defined as the number of months since baseline. For significant group × time interactions, post-hoc pairwise comparisons were performed to examine within-group changes over time using the emmeans package. Estimated marginal means were calculated for each group at baseline and 48-month follow-up, and pairwise contrasts were computed to assess the magnitude and significance of longitudinal change within each group separately.

We used an ANCOVA approach to test whether changes in neurophysiological variables of interest that showed a PD-related change over time also correlated with clinical progression, controlling for its baseline value, change in LEDD, and time to follow-up:

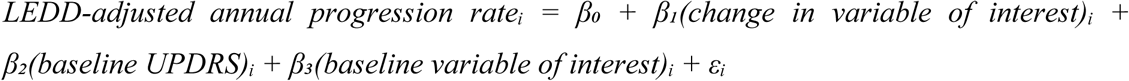

False Discovery Rate (FDR) method was used to control for multiple comparison across all models, unless otherwise specified in the text. Visualization of results was achieved using the *ggseg* package in R.

### Prediction of future symptom progression based on baseline neurophysiological data

Given our results presented below indicating that different aspects of the neuronal signal were differentially associated with clinical symptoms, we next wanted to know to which extent the combination of these features is predictive of future clinical progression of motor symptoms across brain regions and how well this prediction is transferable to a new set of persons with PD. To this end, non-demented patients from our baseline cohort who did not participate in the follow-up MEG recordings but for whom clinical follow-up data was available were included in the validation cohort. Note that here only the total MDS-UPDRS III scores were available.

Because of the expected multicollinearity of the features of the neurophysiological signal across brain regions, it was deemed inappropriate to include them into the same mixed-model. Instead, we used a combination of Principal Component Analysis (PCA) and Partial Least Square Regression (PLS).

First, baseline neurophysiological variables of interest (aperiodic exponent, aperiodic offset, peak alpha frequency and peak beta power) underwent region-wise PCA to reduce dimensionality while preserving regional neurophysiological signatures. For each brain region, the first principal component (PC1) was extracted, accommodating missing data through dynamic feature selection without imputation. I.e., in case of missing values for one of the oscillatory features, the PCA for this region was calculated considering only the remaining three features. This approach created regional summary scores (i.e., PC1) that captured the dominant pattern of spectral variation within each region. The resulting regional PC1 scores served as predictors in a partial least squares regression to predict the LEDD change-adjusted annual MDS-UPDRS III progression rate. Leave-one-out cross-validation was used to determine the optimal number of PLS components. Next, variable importance was assessed using Variable Importance in Projection (VIP) scores, where regions exceeding VIP > 1.0 are generally considered highly predictive. Model performance was evaluated using coefficient of determination (R²) and root mean square error (RMSE) on both training data and then on the independent external validation cohort (details see below) to assess generalization capability.

### Data availability

All custom code used to process the data and run the analysis presented in the paper is available at: https://github.com/JoWld/pd_longitudinal

The full dataset cannot be made publicly available, as the ethical permits for the study do not allow for open data sharing. We have released baseline MEG data used in this analysis in anonymised form.^24,34^ The data is available through the EBRAINS data-sharing platform.^35^ Access to the data requires that the acceptance of the EBRAINS Data Usage Agreement for human data requires all users to create an account and identify themselves using an institutional email address.

## Results

### Longitudinal progression of symptom severity

Complete clinical and MEG data were collected from 28 persons with PD and 31 healthy individuals matched for age and sex. After data collection, the diagnosis in one participant in the PD group was revised to the Parkinsonian type of Multiple System Atrophy (MSA-P) and the participant was subsequently excluded from our analysis. One healthy participant with a positive family history for Essential Tremor displayed postural and kinetic tremor and mild bradykinesia at follow-up (MDS-UPDRS part III 9 points) and was therefore excluded from further analysis.

Please see Table 1 for a summary of demographics and clinical characteristics of the final cohort consisting of 30 healthy individuals and 27 persons with PD (17 with right-lateralized and with 11 left-lateralized symptoms, respectively).

The mean follow-up period was 48.8 ± 7.3 months. In the PD group, motor symptom severity progressed only slightly over the follow-up period with a mean increase in MDS-UPDRS III score of 1.4 ± 8.6 points without reaching statistical significance (Table 1). On an individual level, motor disease progression was highly variable ranging from a reduction by 18 points in MDS-UPDRS III to an increase by 16 points in MDS-UPDRS III (**Supplementary Fig. 2**). Eleven patients (41%) showed a clinically meaningful decline in motor functions defined as an increase in MDS-UPDRS III of more than 4.6 points^36^ whereas nine patients remained stable and seven showed clinically relevant improvement in motor function. Here, it is important to mention that the levodopa equivalent daily dosage increased in all patients with almost double the baseline dose at follow-up on a group level (Table 1). However, the relationship between LEDD adjustments and clinical progression seems to be complex as there was no evidence supporting that change in LEDD alone could explain change in MDS-UPDRS III or its subscores in a linear regression model (total MDS-UPDRS III: *r =* −1.130, *p =* 0.226, **Supplementary Fig. 2**).

General cognitive abilities measured in MoCA remained stable in both groups (Table 1). While three persons with PD and two healthy control participants scored under the cut-off for potential cognitive impairment of 23/24 points in MoCA, none of the recruited participants fulfilled clinical criteria for a diagnosis of dementia.

### PD-related neurophysiological alterations

At baseline, there was evidence for a decreasing effect of PD on peak alpha frequency in bilateral superior temporal (right: *t(27) =* −3.767, *p_FDR_ =* 0.020; left: *t(27) =* −3.294, *p_FDR_ =* 0.023), right transverse temporal (*t(27) =* −4.582, *p_FDR_* = 0.004) region, and right pars triangularis of the inferior frontal gyrus (*t(27) =* −3.711, *p_FDR_* = 0.023) (28% of ROI excluded from analysis for which values were available for < 10 participants per group, **Supplementary Fig. 3**). Further, we identified an increasing effect of PD on peak beta power in a large range of cortical areas accentuated in parietal, temporal and occipital regions (**Figure 3**). Likewise, both the exponent and offset of the aperiodic component were higher in the PD group compared to healthy participants with a similar spatial pattern (**Supplementary Fig. 3**). No evidence for group differences was found for peak beta frequency or peak alpha power.

Within the PD group, peak beta power was negatively correlated with bradykinesia in the right superior parietal region (*t(22) =* −3.250, *p_FDR_* = 0.049), left lateral occipital region (right: *t(22) =* −3.423, *p_FD_*_R_ = 0.044), right fusiform gyrus (*t(22) =* −3.671, *p_FDR_* = 0.044) and left caudal anterior cingulate cortex (*t(22) =* −3.395, *p_FDR_* = 0.044) with the same trend in the right inferior parietal (*t(22) =* −3.089, *p_FDR_* = 0.052) and right lateral occipital regions (*t(22) =* −3.091, *p_FDR_* = 0.052) (**Figure 3**).

### Longitudinal change of aperiodic and periodic components

We first aimed to compare the change in the distribution of peaks in the entire frequency spectrum between 1 and 35 Hz across the whole cortex, i.e., without a priori delineation of the signal into traditional frequency bands (**Figure 1**). From visual inspection, one may appreciate that PD was associated with a progressive reduction of peak frequency in the alpha and low beta frequency ranges over time.

In the analysis by region and frequency band, no evidence for significant group*time interaction in the mixed-effects models for any of the oscillatory neurophysiological variables of interest was found (ROI exclusion rate in the extended alpha band: 37 %). A trend towards an effect of the group*time interaction for peak beta power in the left caudal middle frontal gyrus (*t(51.4) =* 3.146, *p_FDR_ =* 0.09) and left paracentral region (*t(51.5) =* 3.387, *p_FDR_ =* 0.09) did not survive correction for multiple comparison. Since we had a strong a priori hypothesis about PD-related changes in beta power over time, we conducted an exploratory by-group analysis. Here, we did not find evidence for any regional changes in peak beta power in the healthy control group (all *p_FDR_* > 0.05). The PD group, on the other hand, showed an increase in peak beta power in several additional regions encompassing the left pre- and postcentral region, parts of the inferior and posterior middle frontal gyrus, and right temporal lobe and insula (**Figure 4**). Furthermore, there was evidence for a decrease in peak beta power over time in the left lateral occipital region and bilateral cuneus, whereby the raw peak beta power at follow-up was still higher than in the healthy control group (**Supplementary Fig. 4**).

Regarding longitudinal change of the aperiodic component, there was a significant group*time interaction for the exponent of the aperiodic component in the left postcentral (*t(55) =* 3.424, *p_FDR_* = 0.047) and right paracentral regions (*t(55) =* 3.367, *p_FDR_ =* 0.047). In group-wise post-hoc comparison, the PD group exhibited an increase over time in the left postcentral region (*t(26) =* 3.358, *p_FDR_ =* 0.032) while the healthy control group did not display significant change with a general tendency towards a decreasing exponent in the central region (**Figure 2**).

**Figure 2:**
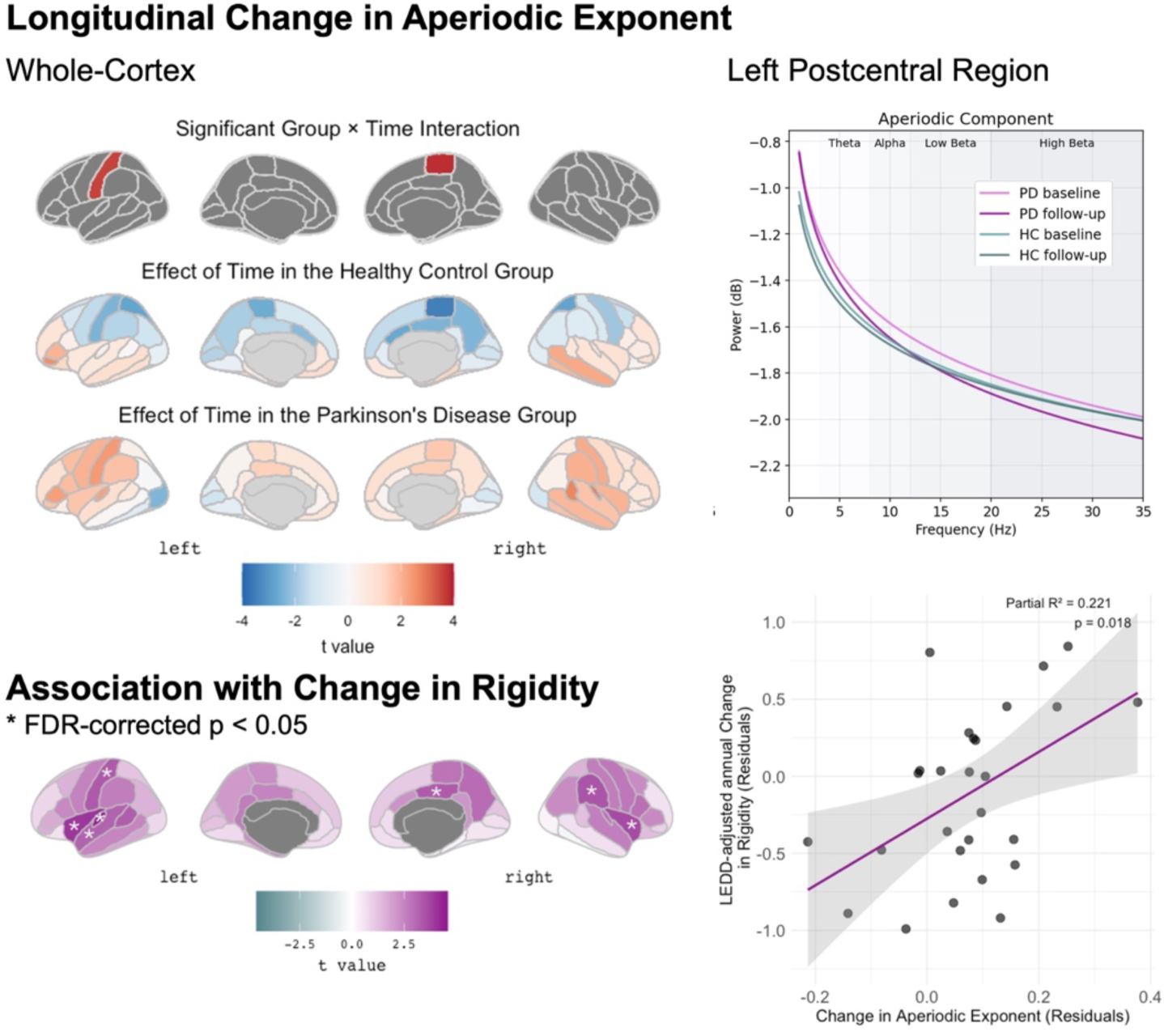
Longitudinal change in the aperiodic exponent and association with change in rigidity across the cortex and in the left postcentral region of interest as an example (right panel). **Left panel: Upper row:** Group x time interaction across brain regions for the aperiodic exponent (upper row), only displaying t statistics for regions with a significant interaction effect defined by p_FDR_ < 0.05. **Lower rows:** Main effects of time per group for illustration of the overall trends over time. Lowest row show illustrating the results of the ANCOVA for the relationship between change in aperiodic exponent and rigidity progression. * indicates regions with for which the ANCOVA model described in the main text resulted in a significant effect with an FDR-corrected p-value < 0.05. Colouring represents the respective t values of this effect. **Right panel: Upper row:** Aperiodic component of the neuronal signal as derived from parametrization by group and time point in the left postcentral region. **Lower row:** Relationship between change in the aperiodic exponent and annual rigidity progression rate. R² in the upper right corner is the partial R², indicating how much of the variance in clinical change is explained by adding change in the aperiodic exponent to the ANCOVA model. The grey area represents the 95%-confidence interval around the fitted regression line in pink.

### Relationship of longitudinal neurophysiological changes with symptom progression

Based on the evidence for a significant PD-related longitudinal change in the aperiodic exponent, we next aimed to explore whether this change related to clinical progression, as measured in MDS-UPDRS III and its subscores.

The ANCOVA model resulted in a significant positive association of the change in exponent of the aperiodic exponent with the annual LEDD-adjusted change in rigidity in the left postcentral gyrus (*t(26) =* 3.445, p_FDR_ = 0.030), right supramarginal gyrus (*t(26) =* 3.544, *p_FDR_ =* 0.030) as well as parts of the left superior temporal lobe and bilateral insula (**Figure 2**). The same trend was found for additional adjacent regions, including bilateral precentral and paracentral, right postcentral, and left paracentral and supramarginal gyrus (all p_FDR_ = 0.068).

For completeness, it is worth mentioning that there was evidence for a significant effect of the *baseline* aperiodic exponent on the annual progression rate of bradykinesia in bilateral precuneus (right: *t(26) =* 3.372, *p_FDR_* = 0.045; left: *t(26) =* 4.593, *p_FDR_ =* 0.009), bilateral superior parietal (right: *t(26) =* 3.489, *p_FDR_ =* 0.045; left: *t(26) =* 3.595, *p_FDR_ =* 0.045), and left paracentral regions (*t(26) =* 3.278, *p_FDR_* = 0.045).

Furthermore, we identified significant positive associations between change in the aperiodic offset and progression of bradykinesia in bilateral entorhinal (right: *t(26) =* 3.103, p_FDR_ = 0.043; left: *t(26) =* 3.453, p_FDR_ = 0.043), parahippocampal (right: *t(26) =* 3.450, p_FDR_ = 0.043; left: *t(26) =* 3.317, p_FDR_ = 0.043), inferior temporal (right: *t(26) =* 3.140, p_FDR_ = 0.043; left: *t(26)* 3.674, p_FDR_ = 0.043), and right fusiform gyri (*t(26) =* 3.258, p_FDR_ = 0.043) (**Supplementary Fig. 5**).

Lastly, we explored whether the negative relationship between peak beta power and severity of bradykinesia found at baseline changed over the follow-up period. Here, we hypothesized that this relationship might weaken over time if it paralleled compensatory mechanisms and their assumed break down with disease progression. To this end, we built a mixed-effects model with bradykinesia as dependent variable, and peak beta power, time, their interaction, and LEDD as fixed effects and subject as a random effect. In this model, there was evidence for a significant peak beta power x time interaction in the right superior parietal region (*t =* 3.011, *p_FDR_ =* 0.019), indicating a significant change in the relationship between peak beta power and bradykinesia over time (**Figure 3**). For post-hoc comparison, we further divided the PD cohort based on whether they progressed in bradykinesia over time (defined by LEDD-adjusted annual change > 0). Here, it became evident that the significant negative association of peak beta power and bradykinesia at baseline vanished in progressors while it remained in the non-progressors (yet, without reaching statistical significance) (**Figure 3**).

**Figure 3:**
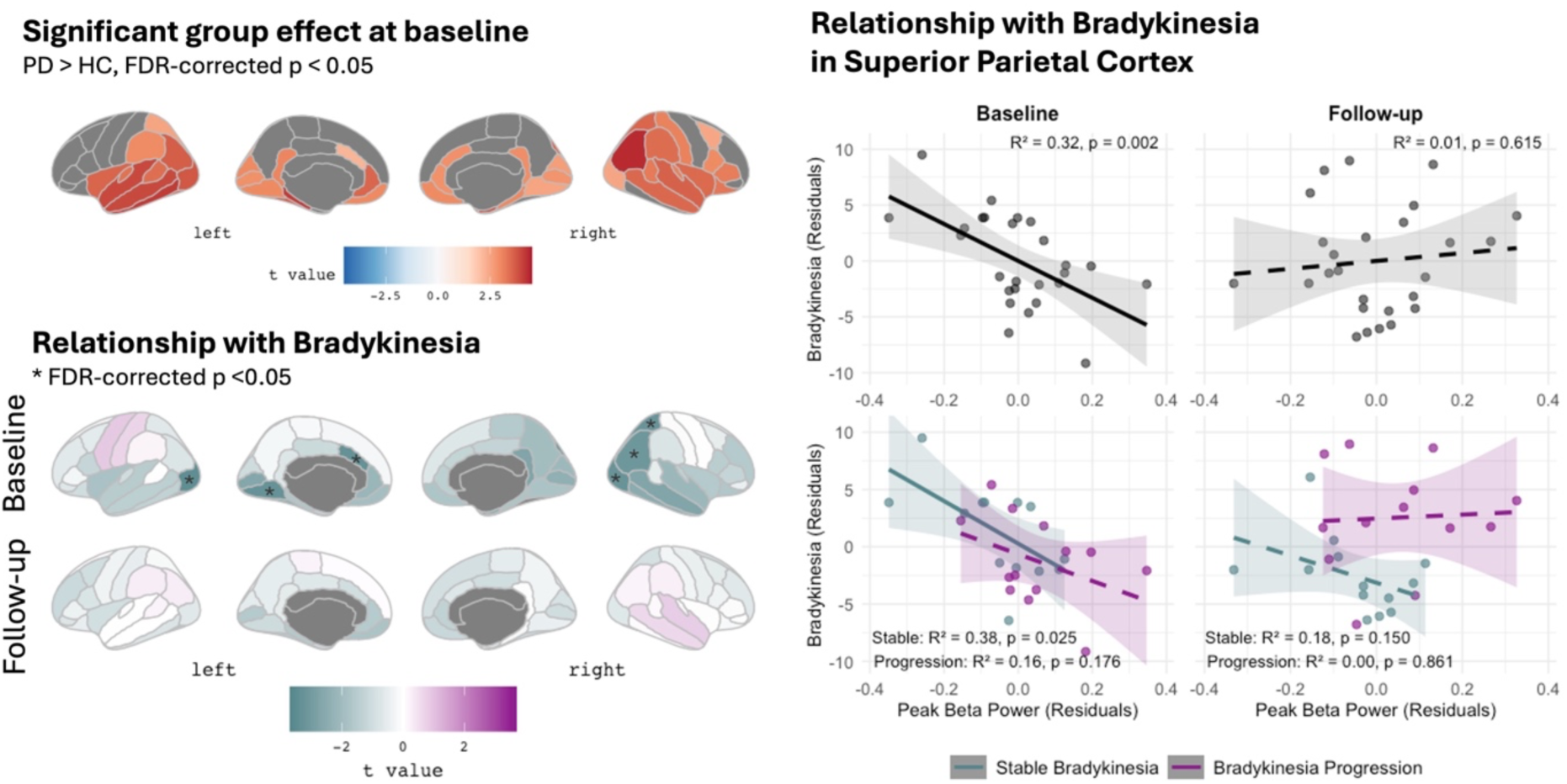
Pattern of PD-related increase in peak beta power and its relationship with bradykinesia at baseline and follow-up. **Right panel: Upper row:** Only showing color-coded t statistics for regions with a significant group effect defined by p_FDR_ < 0.05 across all cortical regions. There was a general trend towards a further increase of peak beta band power over time except for the occipital lobe (Supplementary Fig. 4). **Lower rows:** Results of the regression model for the association between peak beta power and bradykinesia corrected for LEDD and age. * indicating an FDR-corrected p < 0.05. **Right panel:** Exemplary results from the superior parietal region. Dashed lines represent insignificant effects of peak beta power on bradykinesia. Shaded areas show the 95%-confidence interval around the fitted regression lines. In the lower row, patients were post-hoc sub-divided by annual progression rate of bradykinesia with progressors showing a rate > 0, i.e., an increase of bradykinesia severity over time.

### Predicting Symptom Progression from Neurophysiology

We next wanted to explore how much of the longitudinal clinical progression could be explained by simple aperiodic and periodic features of the neuronal signal at baseline. So far, our analysis focused on the effects of one of these features at a time. However, they showed collinearity which is reflected by our results suggestive of associations of several features (peak beta power, aperiodic exponent and offset) with aspects of motor symptoms (bradykinesia and rigidity) and their longitudinal progression, respectively (see **Supplementary Fig. 6** for correlation matrix). Combining all these features into one prediction model is further complicated by the multicollinearity across the 68 parcellated brain regions. Additionally, our PD sample was relatively small resulting in a risk for overfitting. We therefore opted for a combined approach with dimension reduction across neurophysiological features using PCA followed by partial least squares regression of the first component with the LEDD-adjusted annual progression rate of the MDS-UPDRS III score across regions.

Our validation cohort consisted of 18 persons with PD, well-matched to the training cohort in terms of age, sex, baseline motor symptom severity, and LEDD (age: 65.4 ± 10.2 years, *t(43) =* −0.24, *p =* 0.8, baseline MDS-UPDRS III: 19.1 ± 12.1, *t(43) =* 0.37, *p >* 0.9), **Supplementary Table 1**). Paralleling the longitudinal motor symptom progression in our original training sample, the validation cohort did not show significant change in MDS-UPDRS III (*t (17) =* −1.35, *p =* 0.195) while considerably increasing LEDD over time (*t (17) =* −4.24, *p =* 0.001).

In the training cohort, the region-wise PCA resulted in first components (PC1) explaining 62.8 ± 8.5 % of variance across regions (range 44.6 − 80.0%). Partial least squares regression with leave-one-out cross-validation identified two components as optimal for progression rate prediction. The model achieved a moderate effect size on training data *(R² =* 0.365, *RMSE =* 1.676) while external validation revealed reduced generalization (*R² =* 0.195, *RMSE =* 3.674).

Using a permutation test, the observed R² in the validation cohort exceeded 93.1% of values in the null distribution, yielding a non-significant permutation p-value of 0.069 (10,000 permutations). Adding age, sex, and baseline MDS-UPDRS to the model increased performance to 25.0 % in the validation cohort (training: R² = 0.578, RMSE = 1.366, validation: R² = 0.250, *RMSE =* 3.475, *p =* 0.030 (10,000 permutations).

The performance difference between the training and validation cohorts suggests overfitting to the training data that increased further when adding clinical variables, likely reflecting the relatively high-dimensional nature of the neuroimaging input relative to sample size (2.5:1). Still, we can conclude that 19.5 % of variance in motor progression in the independent sample was explained solely by the first component of the PCA constructed from peak beta power, peak alpha frequency and the aperiodic component of the neuronal data, rising to 25 % when adding age, sex and motor symptom severity at baseline.

Next, we explored the differential contributions of brain regions and neurophysiological variables to the overall model. The aperiodic component showed the highest average contribution to regional PC1 (exponent: *mean squared loading =* 0.350 ± 0.049, offset: 0.303 ± 0.055, followed by peak beta power (0.258 ± 0.074), and peak alpha frequency (0.089 ± 0.071). Likewise, aperiodic features contributed most to PC1 in a total of 62 regions with predominantly positive loadings (**Figure 4**). Peak beta power was most influential in the remaining six regions, including negative loadings in bilateral inferior parietal and left superior parietal regions (*max squared loading =* 0.317) and positive loadings in the right fusiform and parahippocampal regions (*max squared loading* = 0.349). Similar patterns emerged when the loadings were weighted by VIP scores derived from the PLS, suggesting that variations in the aperiodic component were also most consistently associated with clinical progression across high-importance brain regions. Bilateral central and superior parietal cortices were identified as the most contributing regions, showing the highest VIP scores and a consistent at least small effect size (*Cohen’s d* > 0.2) for the correlation between the regional PC1 and progression rate at both baseline and follow-up (**Figure 4**).

**Figure 4:**
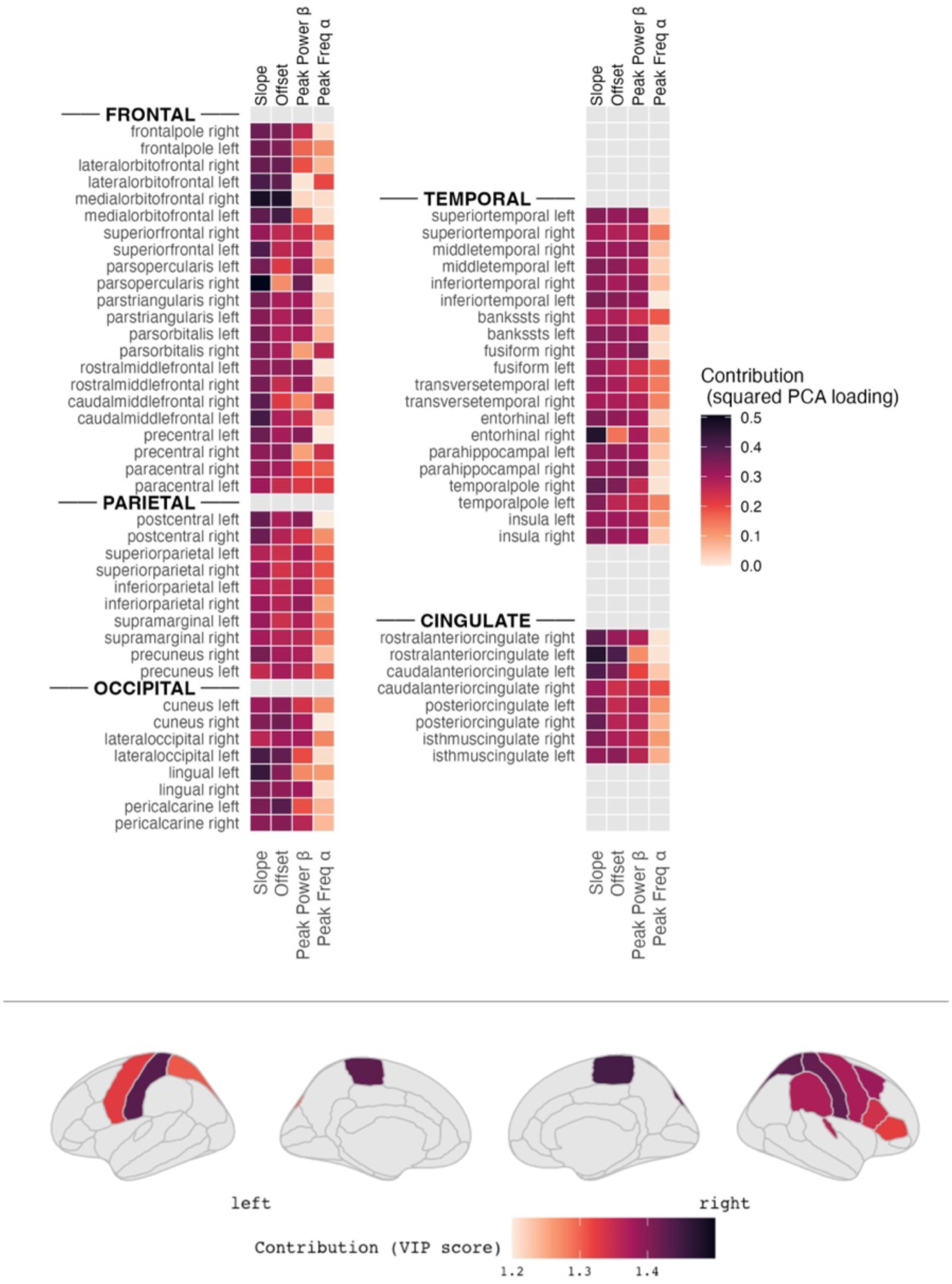
Contributions of power spectrum features and regions to the predictive model. **Upper row**: Mean squared loading of each of the four spectral features (aperiodic exponent, aperiodic offset, peak beta power and peak alpha frequency from left to right) color-coded for each region. **Lower row:** Largest regional contributions to the partial least square regression color-coded by variable importance in projection (VIP), only displaying values for regions for which PC1 and annual progression rate were correlated with at least small effect size (Cohen’s d = 0.2) at both baseline and follow-up. Higher VIP, i.e., darkest colour, representing largest contribution.

## Discussion

### Summary of findings

In this longitudinal study, we analysed the relationship between changes in motor symptoms and changes in whole-cortex neurophysiological activity in a cohort of persons with PD over an average period of four years. Our results show that the exponent of the aperiodic component of the neurophysiological signal in the central sensorimotor region showed a PD-related increase over time that went beyond healthy aging, and longitudinal change of the aperiodic component (represented by its exponent and offset) was associated with progression rates of rigidity and bradykinesia, respectively.

The progressive PD-related alteration of the aperiodic component over time emphasizes the need for including as well as correcting for aperiodic activity when assessing oscillatory brain activity in PD. After removal of the aperiodic component and stringent correction for multiple comparison, we did not find evidence for consistent PD-related longitudinal change in peak power or frequency in the beta or alpha frequency bands in any of the parcellated brain regions. However, at baseline, there were wide-spread temporo-occipito-parietal increases of peak beta power in PD compared with healthy individuals that persisted over time. A negative association between parieto-occipital peak beta power and bradykinesia at baseline changed significantly over time in the right superior parietal cortex, in particular in patients that experienced clinically progressive bradykinesia. Taken together, these findings suggest that increased parietal beta power in early PD may be associated with compensation.

Lastly, we predicted longitudinal clinical symptom progression based on aperiodic (exponent and offset) and periodic features (peak beta power and peak alpha frequency) of the neurophysiological signal. Here, 19.5% of the variance in the clinical progression over time was explained by these basic features of the neuronal signal in an independent validation cohort.

### Limited clinical symptom progression in our cohort

Previous cross-sectional evidence suggests disease-stage-dependent PD-related alterations of beta band activity in the motor cortex in early PD with an increase of beta power in de-novo and early PD that decreases with disease progression, eventually falling below healthy levels.^6,37^ Our results do not support this claim; instead, we found a trend towards a further increase from early to mid-stage PD (at least in the left hemisphere). Here, it is worth noting that, despite a follow-up period of more than four years, our cohort did only exhibit a marginal increase in motor symptoms in on-medication state on a group level with considerable variability between patients. The lack of significant change in oscillatory cortical activity may in part reflect the lack of clinical progression in approximately half of our sample.

Likewise, participants included in our follow-up assessment remained cognitively stable which may contribute to the lack of a relationship of cognitive performance with cortical activation patterns over time. Thus, our findings may not be generalizable to cohorts with progressive cognitive impairment. On the other hand, the fact that our sample remained cognitively stable (NB, according to a cognitive screening test) is an advantage as it supports that the effects seen with motor progression are less likely to be influenced by concomitant cognitive decline.

### The aperiodic component and longitudinal disease progression

The previous longitudinal study of neurophysiological resting state activity in PD estimated relative power in several frequency bands without separating or controlling for the effects of the aperiodic component of the signal from that of the periodic component. In their sample with a follow-up period of up to seven years, “oscillatory slowing” (manifested as an increase in theta power and decrease in alpha power) was progressive^7^ and most pronounced in centro-parieto-temporal regions.^23^ Here, it is worth noting that publications on this cohort averaged either across the entire cortex (in sensor space) or into large subregions roughly corresponding to lobes, respectively. Our analysis approach adds to these prior studies by taking into account and compensating for the aperiodic component and by performing a more fine-grained analysis of the spatial distribution. While we found evidence to support a shift in alpha activity to lower frequencies when averaging across the entire cortex (Figure 1), this was not further substantiated specific regional PD-related patterns of longitudinal decrease in peak alpha frequency (4-12 Hz) when controlling for multiple comparison. In agreement with our findings, Boon et al. found a longitudinal association of *lower* relative beta band power with *worse* motor performance (measured as UPDRS III) with a maximum of this effect located in the temporal, occipital and inferior parietal regions.^7^ However, they attributed the progressing reduction in relative beta power to the pathological shift of the overall signal to lower frequencies. In line with this interpretation, a longitudinal increase in theta power in the PD group was associated with more severe motor symptoms across several brain regions sparing the frontal lobe (in sensor space) in the earlier paper reporting longitudinal findings in the same cohort.^23^ These previous results indicating a general progressing oscillatory slowing in PD should be interpreted with caution given our results of a progressive alteration of the aperiodic component. A steeper slope of the aperiodic component could lead to a similar pattern of change even if the underlying oscillatory activity in the theta and alpha / beta frequency ranges, respectively, remained unchanged. Our results offer a new perspective to neurophysiological changes in PD as they emphasize that the most prominent PD-related longitudinal change in resting state neuronal activity is a steepening of the aperiodic slope or *aperiodic slowing.* Furthermore, we found that longitudinal change in the aperiodic component was associated with clinical progression of bradykinesia and rigidity. Our findings (a relationship of change in the aperiodic and change in clinical symptom severity, but not with absolute values at each timepoint) is in line with recent work on aperiodic activity in the subthalamic nucleus where changes in the aperiodic slope after levodopa intake correlated with motor symptom alleviation.^38^ Within-subject trajectories of the aperiodic component may have larger predictive value of clinical status than static, cross-sectional measures, potentially due to high inter-individual variability.

### Cortical beta and compensation

As described in the introduction, the relationship between cortical beta and motor symptoms in PD seems to be more complex than in the subcortex where most studies identified a clear link between higher beta power and more severe bradykinesia or rigidity (see ^39^ for review). Our findings add to the existing cross-sectional evidence for paradoxical effects of PD itself and its treatment on cortical beta activity, challenging the interpretation of beta as universally antikinetic. Interestingly though, studies on cortical beta (oscillations and bursts) still report the strongest relationship with bradykinesia above other motor symptoms (if any)^10,22^, indicating shared underlying mechanisms. Here, it is also worth noting that negative relationships between cortical beta power and motor symptoms have occasionally been reported before both cross-sectionally^40^ and longitudinally.^7^ However, in contrast to previous interpretations, we do not consider this relationship to be fully explainable with an oscillatory slowing effect as beta power was *increased* compared to healthy individuals across many brain regions at both time points in our study. Instead, we argue that parieto-temporo-occipital beta increases may be associated with compensation. Indeed, the temporo-parieto-occipital spatial distribution of this relationship at baseline is in line with recent work from Wiesman et al. reporting (aperiodic) neurophysiological slowing with a similar spatial pattern was associated with less motor symptoms.^16^ Furthermore, roles of the parietal cortex, the posterior cingulate, and occipito-temporal areas for compensation in PD have been postulated before in several fMRI studies.^41–43^ Regarding beta activity, elevated beta coherence in the superior parietal cortex as well as between the supplementary motor cortex and the primary motor cortex has previously been discussed as compensatory mechanisms.^44,45^ Different roles of beta activity in different cortical regions may be explained by a proposed functional polymorphism of beta activity.^46^ While a direct link between beta oscillations and the dopamine system has recently been further substantiated in humans^47^, there are several sources of dopaminergic modulation to the cortex with beta activity in the basal ganglia explaining as little as 20% of the variance of the cortical beta signal.^46^

That the relationship weakened over time paralleling clinical progression in bradykinesia while it remained intact in clinically stable patients further supports the interpretation as potentially associated with compensation. Still, lacking information about the individual degree of subcortical neurodegeneration (and its progression over time), this interpretation remains speculative.

### Predicting future symptoms from cortical neurophysiological data

Most cross-sectional predictive MEG/EEG studies in PD so far focused on models for diagnosis or current symptoms rather than prediction of future motor symptoms. Prediction of future clinical progression in PD based on M/EEG data has been most successful in foreseeing cognitive decline.^48–51^

In our cohort, almost 20% of the longitudinal variation in motor symptom progression could be predicted from baseline resting-state MEG recordings in an early motor disease stage in an independent cohort. Adding basic demographics and motor performance at baseline to the model increased the predictive value to 25% with an average prediction error of 3.5 points which is below the threshold that is considered meaningful clinical change.^36^ Hence, our results outperform the few previous neuroimaging studies with external validation of continuous models for future motor progression reporting R² values below 10%.^52^ While the accuracy of our model is still far from clinically useful, these results still offer valuable new insights into the relationship between periodic and aperiodic cortical activity and disease mechanisms of PD. It is noteworthy that the aperiodic component tended to be more influential in predicting future motor symptom progression than oscillatory activity at rest. Furthermore, negative loadings of peak beta power as the most influential feature in the right superior and inferior parietal cortex provide additional support for the potential association with compensation discussed above.

### Limitations and Future Directions

First and foremost, the relatively small sample size of our study may have limited statistical power to detect subtle effects and increased the risk of type II errors, potentially leading to an underestimation of true associations between neurophysiological measures and clinical outcomes. However, the use of an independent validation cohort in the predictive part of our study strengthens our findings by demonstrating that the observed relationships can be replicated in a separate sample, thereby reducing concerns about overfitting and chance findings.

We cannot exclude effects of dopamine-replacement therapy on our results because participants were recorded in on-medication state only. The majority of previous studies did not find a significant impact of DRT on cortical activity in resting-state^10,15,53,54^, but evidence is mixed with some studies pointing towards a levodopa-induced increase in beta power localized to the motor cortex.^8^ Likewise, evidence whether or not the aperiodic component is influenced by dopaminergic medication is mixed. However, recent studies with slightly larger sample sizes comparing on- and off-medication states suggest no impact of dopamine replacement therapy on aperiodic slope or offset.^20^

In addition, variable times to follow-up might have influenced our results. We accounted for these limitations by interpreting LEDD-adjusted annual clinical change rates in our longitudinal analysis. Still, a linear approach to calculating progression rates may be an oversimplification of the real-life complex and variable clinical trajectories in PD.

Beyond effects of DRT and follow-up period, the lack of significant clinical progression on a group-level is likely in part explained by attrition bias, such as that the least and most severely affected individuals got lost to follow-up. While we can confirm that this was the case in the PD group, we can only assume similar effects in the healthy control group it since we do not have any clinical follow-up data on healthy individuals.

Although it is a strength of our approach that the persons with PD included in the validation were recorded with an identical MEG and MRI protocol at baseline and clinically assessed by the same neurologists, we acknowledge that the validation cohort did not perfectly match the training cohort in terms of disease progression. While the annual progression slopes of the two cohorts were almost identical (**Supplementary Material**), the substantial discrepancy between training and validation performance of our PLS model still indicates limited generalizability of the current model to new patients. These findings thereby do not support clinical utility of our approach to derive personalized predictions for individual patients. They highlight the challenge of reliable biomarker discovery in small neuroimaging cohorts and suggest that either larger sample sizes or alternative analytical approaches may be necessary to achieve clinically meaningful prediction accuracy. Given previous promising results in cross-sectional cohorts, our results still encourage future work including more sophisticated neurophysiological measures, e.g., functional connectivity and complementary non-oscillatory features such as beta bursts.^10,20,22,53,56–58^ The identified spatial pattern of predictive brain regions and neurophysiological measures, while biologically plausible, requires validation in larger external cohorts. Optimally, these new patients would be assessed at multiple time points and with even longer follow-up periods in the future to capture individual trajectories of patients with both slow and fast disease progression.

Our results let us to the hypothesis that some of the PD-related beta power increases located in parieto-temporo-occipital regions may be associated with compensation. So far, we focused solely on resting-state activity. Given that we collected data from the same individuals during active and passive motor tasks, we will next test this hypothesis on task-related activity in relation to individual task performance.

Supporting endogenous compensatory mechanisms to delay or reverse clinical disease progression may be a disease-modifying treatment approach. As the cortex is primarily spared from PD-causing pathology, it seems promising to enhance cortical activity patterns associated with compensatory efforts to support the brain’s inherent ability to cope with or eventually overcome neurodegeneration. This potential long-term application of our results requires replication and external validation of our findings.

### Conclusions

This longitudinal study demonstrates that the most prominent PD-related change in resting-state neurophysiological activity is progressive alteration of the aperiodic component, a feature that significantly correlates with clinical progression of bradykinesia and rigidity. Our findings emphasize the critical importance of including and controlling for the aperiodic component when examining neuronal brain activity in PD.

While we found widespread increases in parieto-temporo-occipital beta power in persons with PD compared with healthy individuals, the negative association between parietal beta power and bradykinesia, particularly in clinically stable patients, suggests an association with compensatory mechanisms.

The ability to predict approximately 20% of the clinical variability in longitudinal motor progression using basic resting-state neurophysiological features derived from short and non-invasive MEG recordings warrants future investigations into their predictive value and the underlying relationship with neurodegenerative and compensatory processes.

## Acknowledgements

The authors thank Niklas Edvall and Karolina af Edholm for assistance with parts of the data collection, and Pascal Helson and Alex Wiesman for their thoughtful feedback discussing the results. Assistance by AI in the form of a Large Language Model tool (Claude, Anthropic) was used for analysis code review and language improvements of the manuscript.

## References

1. Albin RL, Young AB, Penney JB. The functional anatomy of basal ganglia disorders. Trends Neurosci. 1989;12(10):366–375. doi:10.1016/0166-2236(89)90074-x

2. Bologna M, Paparella G, Fasano A, Hallett M, Berardelli A. Evolving concepts on bradykinesia. Brain. 2020;143(3):727–750. doi:10.1093/brain/awz344

3. Kühn AA, Williams D, Kupsch A, et al. Event-related beta desynchronization in human subthalamic nucleus correlates with motor performance. Brain. Published online 2004. doi:10.1093/brain/awh106

4. Little S, Brown P. The functional role of beta oscillations in Parkinson’s disease. Parkinsonism and Related Disorders. Published online 2014. doi:10.1016/S1353-8020(13)70013-0

5. Weinberger M, Mahant N, Hutchison WD, et al. Beta oscillatory activity in the subthalamic nucleus and its relation to dopaminergic response in Parkinson’s disease. J Neurophysiol. 2006;96(6):3248–3256. doi:10.1152/jn.00697.2006

6. Heinrichs-Graham E, Kurz MJ, Becker KM, Santamaria PM, Gendelman HE, Wilson TW. Hypersynchrony despite pathologically reduced beta oscillations in patients with Parkinson’s disease: a pharmaco-magnetoencephalography study. J Neurophysiol. 2014;112(7):1739–1747. doi:10.1152/jn.00383.2014

7. Boon LI, Hillebrand A, Schoonheim MM, Twisk JW, Stam CJ, Berendse HW. Cortical and Subcortical Changes in MEG Activity Reflect Parkinson’s Progression over a Period of 7 Years. Brain Topogr. 2023;36(4):566–580. doi:10.1007/s10548-023-00965-w

8. Cao C, Li D, Zhan S, Zhang C, Sun B, Litvak V. L-dopa treatment increases oscillatory power in the motor cortex of Parkinson’s disease patients. NeuroImage: Clinical. 2020;26. doi:10.1016/j.nicl.2020.102255

9. Melgari JM, Curcio G, Mastrolilli F, et al. Alpha and beta EEG power reflects L-dopa acute administration in parkinsonian patients. Front Aging Neurosci. 2014;6:302. doi:10.3389/fnagi.2014.00302

10. Vinding MC, Tsitsi P, Waldthaler J, et al. Reduction of spontaneous cortical beta bursts in Parkinson’s disease is linked to symptom severity. Brain Communications. 2020;2(1). doi:10.1093/braincomms/fcaa052

11. Stoffers D, Bosboom JLW, Deijen JB, Wolters EC, Berendse HW, Stam CJ. Slowing of oscillatory brain activity is a stable characteristic of Parkinson’s disease without dementia. Brain. 2007;130(Pt 7):1847–1860. doi:10.1093/brain/awm034

12. Bosboom JLW, Stoffers D, Stam CJ, et al. Resting state oscillatory brain dynamics in Parkinson’s disease: An MEG study. Clinical Neurophysiology. 2006;117(11). doi:10.1016/j.clinph.2006.06.720

13. Olde Dubbelink KTE, Hillebrand A, Stoffers D, et al. Disrupted brain network topology in Parkinson’s disease: A longitudinal magnetoencephalography study. Brain. 2014;137(1). doi:10.1093/brain/awt316

14. Olde Dubbelink KTE, Hillebrand A, Stoffers D, et al. Disrupted brain network topology in Parkinson’s disease: a longitudinal magnetoencephalography study. Brain. 2014;137(Pt 1):197–207. doi:10.1093/brain/awt316

15. Stoffers D, Bosboom JLW, Wolters EC, Stam CJ, Berendse HW. Dopaminergic modulation of cortico-cortical functional connectivity in Parkinson’s disease: An MEG study. Experimental Neurology. 2008;213(1). doi:10.1016/j.expneurol.2008.05.021

16. Wiesman AI, da Silva Castanheira J, Degroot C, et al. Adverse and compensatory neurophysiological slowing in Parkinson’s disease. Prog Neurobiol. 2023;231:102538. doi:10.1016/j.pneurobio.2023.102538

17. Donoghue T, Haller M, Peterson EJ, et al. Parameterizing neural power spectra into periodic and aperiodic components. Nat Neurosci. 2020;23(12):1655–1665. doi:10.1038/s41593-020-00744-x

18. Gao R, Peterson EJ, Voytek B. Inferring synaptic excitation/inhibition balance from field potentials. NeuroImage. 2017;158:70–78. doi:10.1016/j.neuroimage.2017.06.078

19. Donoghue T, Schaworonkow N, Voytek B. Methodological Considerations for Studying Neural Oscillations. Eur J Neurosci. 2022;55(11-12):3502–3527. doi:10.1111/ejn.15361

20. Helson P, Lundqvist D, Svenningsson P, Vinding MC, Kumar A. Cortex-wide topography of 1/f-exponent in Parkinson’s disease. NPJ Parkinsons Dis. 2023;9(1):109. doi:10.1038/s41531-023-00553-6

21. Monchy N, Modolo J, Houvenaghel JF, Voytek B, Duprez J. Changes in electrophysiological aperiodic activity during cognitive control in Parkinson’s disease. Brain Communications. 2024;6(5):fcae306. doi:10.1093/braincomms/fcae306

22. Vinding MC, Waldthaler J, Eriksson A, et al. Oscillatory and non-oscillatory features of the magnetoencephalic sensorimotor rhythm in Parkinson’s disease. NPJ Parkinsons Dis. 2024;10(1):51. doi:10.1038/s41531-024-00669-3

23. Olde Dubbelink KTE, Stoffers D, Deijen JB, Twisk JWR, Stam CJ, Berendse HW. Cognitive decline in Parkinson’s disease is associated with slowing of resting-state brain activity: A longitudinal study. Neurobiology of Aging. 2013;34(2). doi:10.1016/j.neurobiolaging.2012.02.029

24. Vinding MC, Eriksson A, Comarovschii I, et al. The Swedish National Facility for Magnetoencephalography Parkinson’s disease dataset. Sci Data. 2024;11(1):150. doi:10.1038/s41597-024-02987-w

25. Goetz CG, Tilley BC, Shaftman SR, et al. Movement Disorder Society-sponsored revision of the Unified Parkinson’s Disease Rating Scale (MDS-UPDRS): Scale presentation and clinimetric testing results: MDS-UPDRS: Clinimetric Assessment. Mov Disord. 2008;23(15):2129–2170. doi:10.1002/mds.22340

26. Nasreddine ZS, Phillips NA, Bédirian V, et al. The Montreal Cognitive Assessment, MoCA: A brief screening tool for mild cognitive impairment. Journal of the American Geriatrics Society. Published online 2005. doi:10.1111/j.1532-5415.2005.53221.x

27. Tomlinson CL, Stowe R, Patel S, Rick C, Gray R, Clarke CE. Systematic review of levodopa dose equivalency reporting in Parkinson’s disease. Movement Disorders. Published online 2010. doi:10.1002/mds.23429

28. Taulu S, Simola J. Spatiotemporal signal space separation method for rejecting nearby interference in MEG measurements. Physics in Medicine and Biology. Published online 2006. doi:10.1088/0031-9155/51/7/008

29. Gramfort A, Luessi M, Larson E, et al. MEG and EEG data analysis with MNE-Python. Frontiers in Neuroscience. Published online 2013. doi:10.3389/fnins.2013.00267

30. Hyvarinen A. Fast and robust fixed-point algorithms for independent component analysis. IEEE Transactions on Neural Networks. 1999;10(3):626–634. doi:10.1109/72.761722

31. Desikan RS, Ségonne F, Fischl B, et al. An automated labeling system for subdividing the human cerebral cortex on MRI scans into gyral based regions of interest. NeuroImage. 2006;31(3):968–980. doi:10.1016/j.neuroimage.2006.01.021

32. Dale AM, Liu AK, Fischl BR, et al. Dynamic statistical parametric mapping: combining fMRI and MEG for high-resolution imaging of cortical activity. Neuron. 2000;26(1):55–67. doi:10.1016/s0896-6273(00)81138-1

33. Bates D, Mächler M, Bolker B, Walker S. Fitting Linear Mixed-Effects Models Using lme4. Journal of Statistical Software. 2015;67:1–48. doi:10.18637/jss.v067.i01

34. Vinding MC, Oostenveld R. Sharing individualised template MRI data for MEG source reconstruction: A solution for open data while keeping subject confidentiality. NeuroImage. 2022;254:119165. doi:10.1016/j.neuroimage.2022.119165

35. Vinding MC, Eriksson A, Comarovschii I, et al. The Swedish National Facility for Magnetoencephalography Parkinson’s Disease Dataset (v1.0). Published online 2023. doi:10.25493/NMD2-2FW

36. Horváth K, Aschermann Z, Ács P, et al. Minimal clinically important difference on the Motor Examination part of MDS-UPDRS. Parkinsonism Relat Disord. 2015;21(12):1421–1426. doi:10.1016/j.parkreldis.2015.10.006

37. Vardy AN, van Wegen EEH, Kwakkel G, Berendse HW, Beek PJ, Daffertshofer A. Slowing of M1 activity in Parkinson’s disease during rest and movement--an MEG study. Clin Neurophysiol. 2011;122(4):789–795. doi:10.1016/j.clinph.2010.10.034

38. Sayfulina K, Filyushkina V, Usova S, et al. Periodic and Aperiodic Components of Subthalamic Nucleus Activity Reflect Different Aspects of Motor Impairment in Parkinson’s Disease. Eur J Neurosci. 2025;61(1):e16648. doi:10.1111/ejn.16648

39. van Wijk BCM, de Bie RMA, Beudel M. A systematic review of local field potential physiomarkers in Parkinson’s disease: from clinical correlations to adaptive deep brain stimulation algorithms. J Neurol. 2023;270(2):1162–1177. doi:10.1007/s00415-022-11388-1

40. Miladinović A, Ajčević M, Busan P, et al. EEG changes and motor deficits in Parkinson’s disease patients: Correlation of motor scales and EEG power bands. Procedia Computer Science. 2021;192:2616–2623. doi:10.1016/j.procs.2021.09.031

41. Sarasso E, Gardoni A, Zenere L, et al. Neural correlates of bradykinesia in Parkinson’s disease: a kinematic and functional MRI study. NPJ Parkinsons Dis. 2024;10(1):167. doi:10.1038/s41531-024-00783-2

42. Johansson ME, Toni I, Kessels RPC, Bloem BR, Helmich RC. Clinical severity in Parkinson’s disease is determined by decline in cortical compensation. Brain. 2024;147(3):871–886. doi:10.1093/brain/awad325

43. van Nuenen BFL, Helmich RC, Buenen N, van de Warrenburg BPC, Bloem BR, Toni I. Compensatory activity in the extrastriate body area of Parkinson’s disease patients. J Neurosci. 2012;32(28):9546–9553. doi:10.1523/JNEUROSCI.0335-12.2012

44. Sparks H, Cross KA, Choi JW, et al. Dorsal visual stream is preferentially engaged during externally guided action selection in Parkinson Disease. Clin Neurophysiol. 2022;136:237–246. doi:10.1016/j.clinph.2021.11.077

45. Pollok B, Kamp D, Butz M, et al. Increased SMA–M1 coherence in Parkinson’s disease — Pathophysiology or compensation? Experimental Neurology. 2013;247:178–181. doi:10.1016/j.expneurol.2013.04.013

46. Jenkinson N, Brown P. New insights into the relationship between dopamine, beta oscillations and motor function. Trends in Neurosciences. Published online 2011. doi:10.1016/j.tins.2011.09.003

47. Chikermane M, Weerdmeester L, Rajamani N, et al. Cortical beta oscillations map to shared brain networks modulated by dopamine. eLife. 2024;13. doi:10.7554/eLife.97184.2

48. Olde Dubbelink KTE, Hillebrand A, Twisk JWR, et al. Predicting dementia in Parkinson disease by combining neurophysiologic and cognitive markers. Neurology. 2014;82(3):263–270. doi:10.1212/WNL.0000000000000034

49. Bertrand JA, McIntosh AR, Postuma RB, et al. Brain Connectivity Alterations Are Associated with the Development of Dementia in Parkinson’s Disease. Brain Connect. 2016;6(3):216–224. doi:10.1089/brain.2015.0390

50. Cozac VV, Chaturvedi M, Hatz F, Meyer A, Fuhr P, Gschwandtner U. Increase of EEG Spectral Theta Power Indicates Higher Risk of the Development of Severe Cognitive Decline in Parkinson’s Disease after 3 Years. Front Aging Neurosci. 2016;8:284. doi:10.3389/fnagi.2016.00284

51. Kozak VV, Chaturvedi M, Gschwandtner U, et al. EEG Slowing and Axial Motor Impairment Are Independent Predictors of Cognitive Worsening in a Three-Year Cohort of Patients With Parkinson’s Disease. Front Aging Neurosci. 2020;12:171. doi:10.3389/fnagi.2020.00171

52. Zeighami Y, Fereshtehnejad SM, Dadar M, Collins DL, Postuma RB, Dagher A. Assessment of a prognostic MRI biomarker in early de novo Parkinson’s disease. NeuroImage: Clinical. 2019;24:101986. doi:10.1016/j.nicl.2019.101986

53. McKeown DJ, Jones M, Pihl C, et al. Medication-invariant resting aperiodic and periodic neural activity in Parkinson’s disease. Psychophysiology. 2024;61(4):e14478. doi:10.1111/psyp.14478

54. George JS, Strunk J, Mak-McCully R, Houser M, Poizner H, Aron AR. Dopaminergic therapy in Parkinson’s disease decreases cortical beta band coherence in the resting state and increases cortical beta band power during executive control. Neuroimage Clin. 2013;3:261–270. doi:10.1016/j.nicl.2013.07.013

55. Wang Z, Mo Y, Sun Y, et al. Separating the aperiodic and periodic components of neural activity in Parkinson’s disease. Eur J Neurosci. 2022;56(6):4889–4900. doi:10.1111/ejn.15774

56. Olde Dubbelink KTE, Stoffers D, Deijen JB, et al. Resting-state functional connectivity as a marker of disease progression in Parkinson’s disease: A longitudinal MEG study. NeuroImage: Clinical. 2013;2(1). doi:10.1016/j.nicl.2013.04.003

57. Pauls KAM, Korsun O, Nenonen J, et al. Cortical beta burst dynamics are altered in Parkinson’s disease but normalized by deep brain stimulation. Neuroimage. 2022;257:119308. doi:10.1016/j.neuroimage.2022.119308

58. Luo CY, Song W, Chen Q, et al. Reduced functional connectivity in early-stage drug-naive Parkinson’s disease: A resting-state fMRI study. Neurobiology of Aging. Published online 2014. doi:10.1016/j.neurobiolaging.2013.08.018

