## Supplementary Material for "Magnetoencephalography-based prediction of longitudinal symptom progression in Parkinson’s disease"

**MEG-based prediction of longitudinal symptom progression in Parkinson's disease: periodic and aperiodic components**

**Supplementary Methods**

**Inclusion criteria:**

- No history, signs or symptoms indicating any (healthy group) or other (PD group) neurological disorders according to a physical and neurological examination
- PD group: diagnosis of idiopathic Parkinson’s disease according to the UK Parkinson’s Disease Society Brain Bank Diagnostic Criteria with Hoehn and Yahr stages 1–3

**Exclusion criteria:**

- diagnosis of major depression, dementia, history or presence of schizophrenia, bipolar disorder, epilepsy or history of alcoholism or drug addiction according to the Diagnostic and Statistical Manual of Mental Disorders V
- metal implants affecting the MEG recordings

**MDS-UPDRS III subscale calculation**

Rigidity: items 3.3a - e

Bradykinesia: items 3.2, 3.4 – 3.8, 3.14

Tremor: items 3.15 – 3.18

Axial Symptoms: items 3.1, 3.9 – 3.13

**Supplementary Results**

**Results for alpha and theta bands separately**

When considering alpha (8-12 Hz) and theta (4-8 Hz) frequency ranges separately, there was a significant group effect at baseline in bilateral superior temporal (right: t = -3.77, p_FDR_ = 0.02, left: t = -3.39, p_FDR_ = 0.02), left transverse temporal regions (t = -3.71, p_FDR_ = 0.02) and right pars triangularis of the inferior frontal gyrus (t = --4.58, p_FDR_ = 0.003). Theta frequency was significantly different between groups in bilateral medial orbitofrontal regions (right: t = 3.90, p_FDR_ = 0.03, left: t = 3.38, p_FDR_ = 0.03). No significant group differences for peak alpha or theta power and associations with motor or cognitive symptoms at baseline were found. Please note that 63% of regions were excluded for theta band based on N < 5 values per group available (this number increasing to 100 % when using a N = 10 threshold).

In the longitudinal analysis, no significant group x time point interactions emerged for alpha or theta power nor frequency.

**Supplementary Figures**

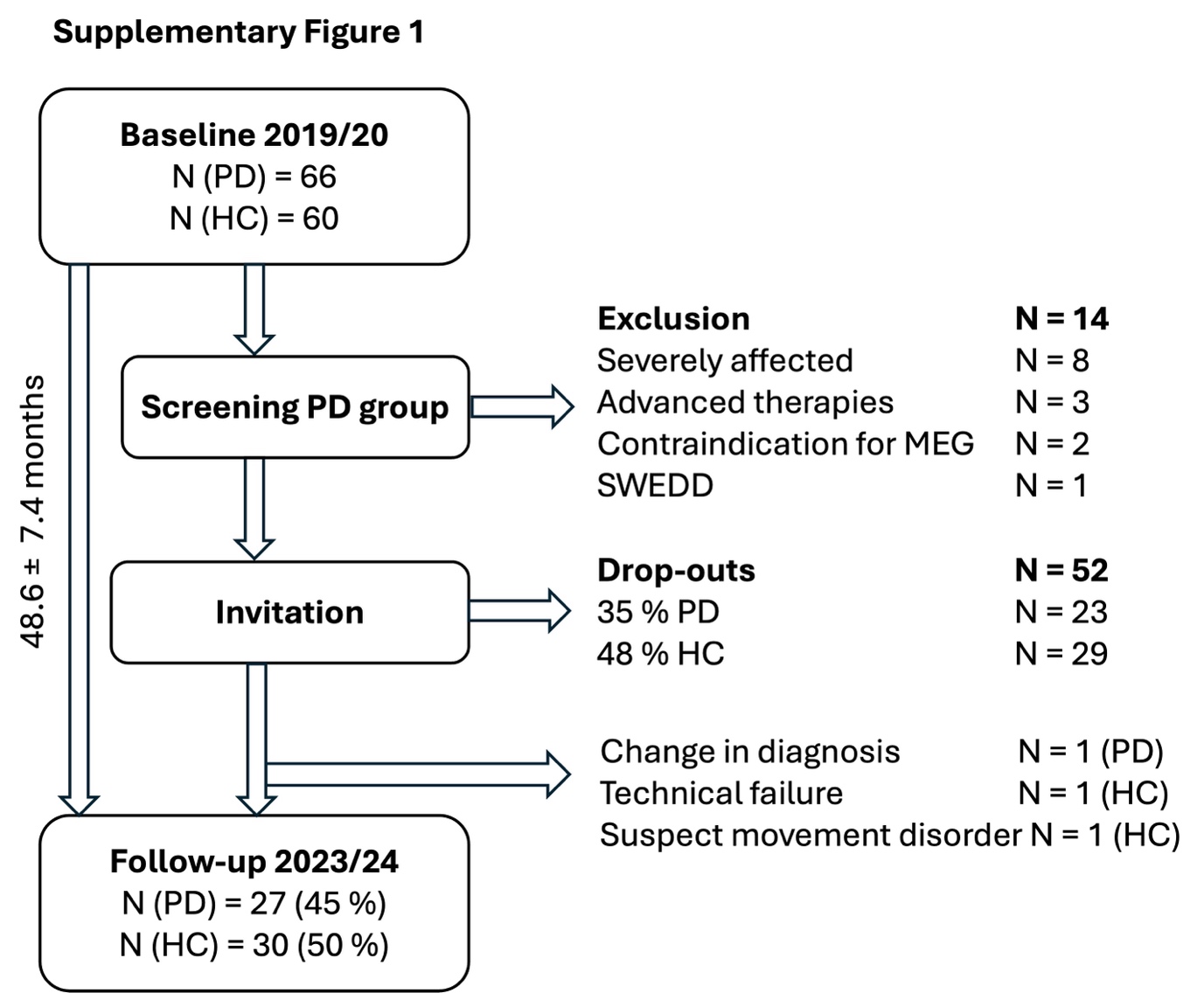

**Supplementary Figure 1: Flowchart of recruitment procedure at follow-up.**

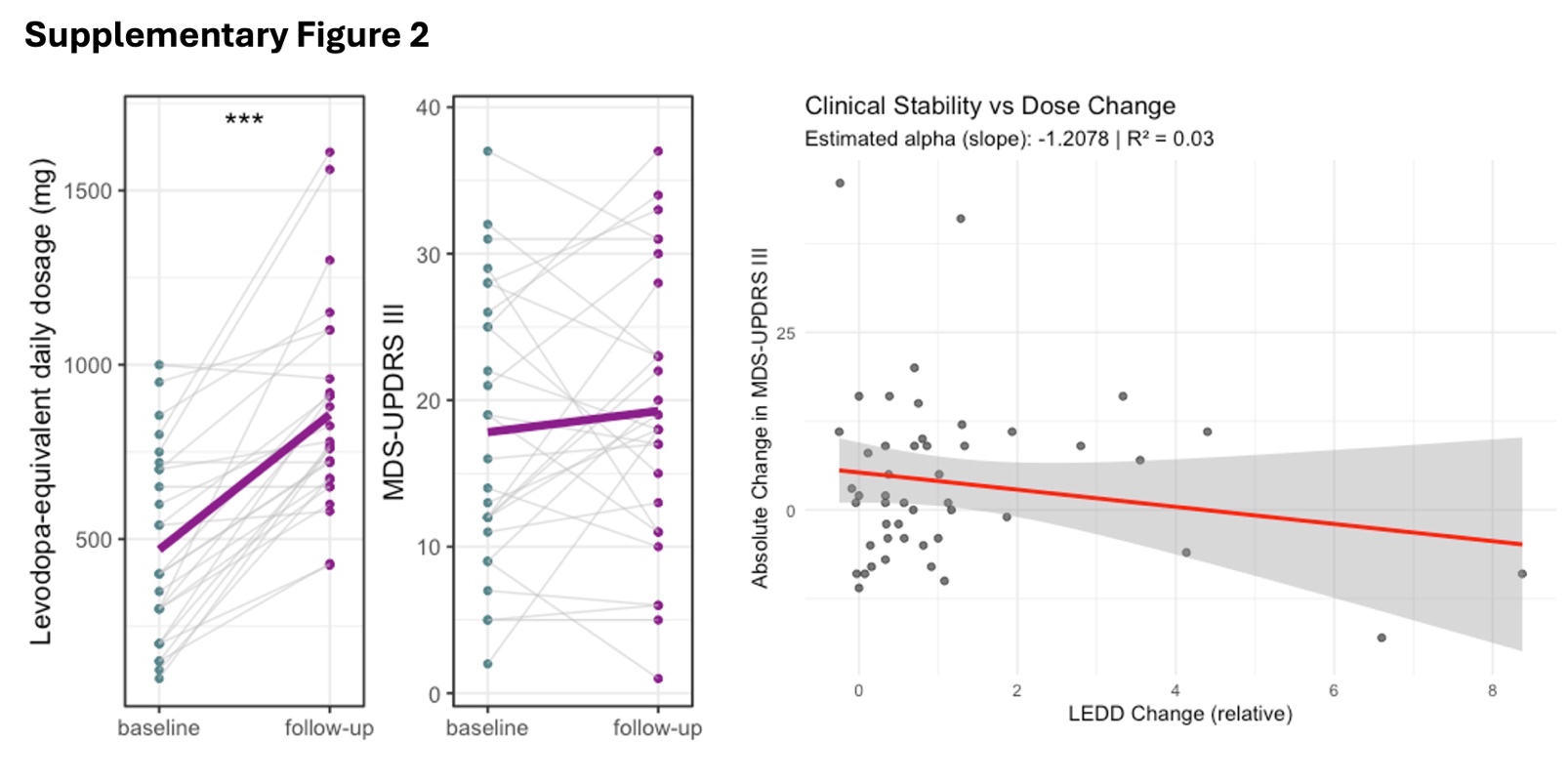
**Supplementary Figure 2: Longitudinal change in medication and motor symptoms. Left:** Individual change in levodopa-equivalent daily dosage (LEDD) and MDS-UPDRS III total score with each dot representing one participant. Within-participant change represented by grey connecting lines, the bold magenta line shows the mean group-level change (N = 27). **Right:** Relationship between the change in MDS-UPDRS and change in LEDD over time. All participants from training and validation cohort included in this graph.

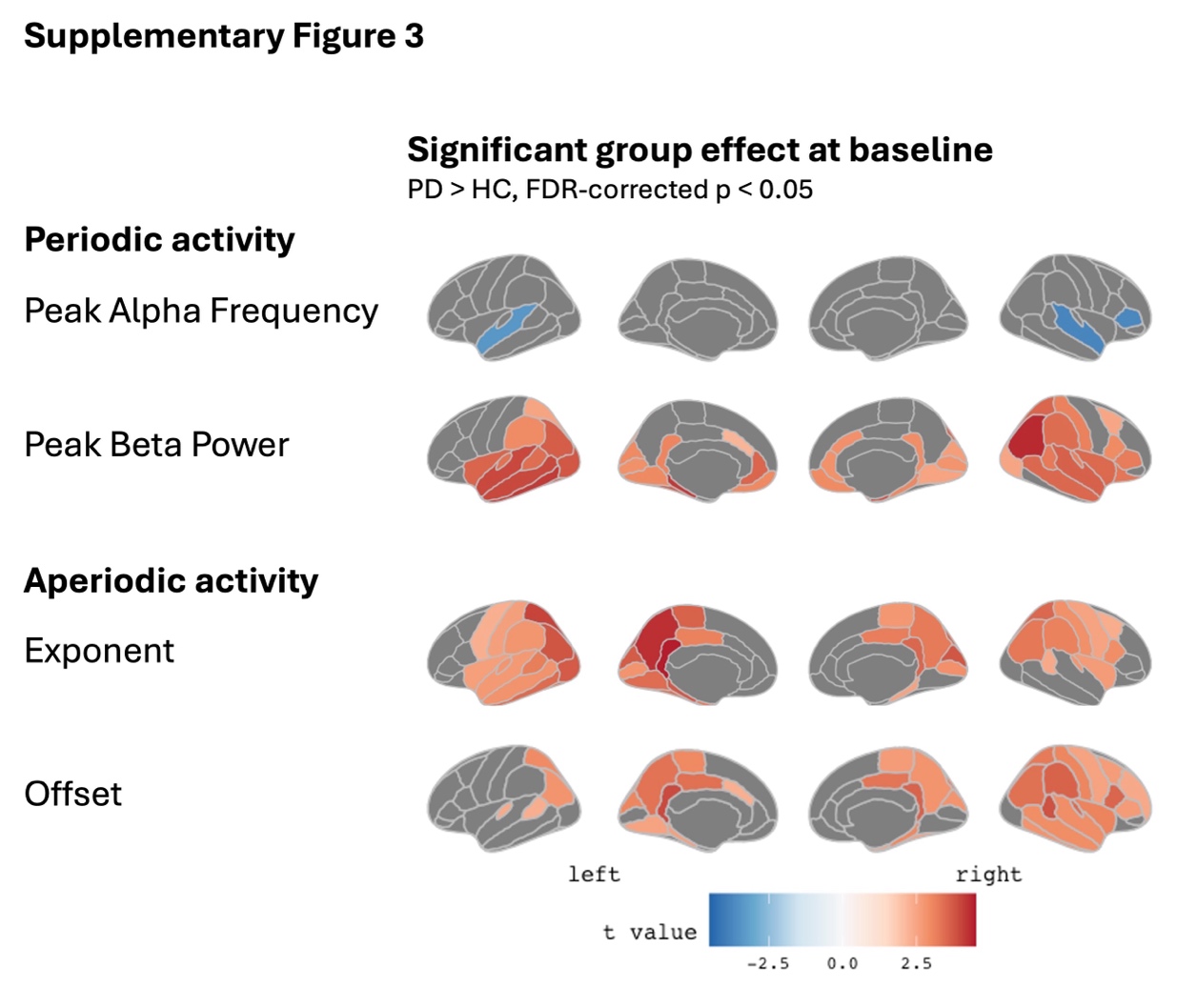

**Supplementary Figure 3: Spatial patterns of PD-related changes at baseline.** Only showing color-coded t statistics for regions with a significant group effect defined by p_FDR_ < 0.05 across all cortical regions.

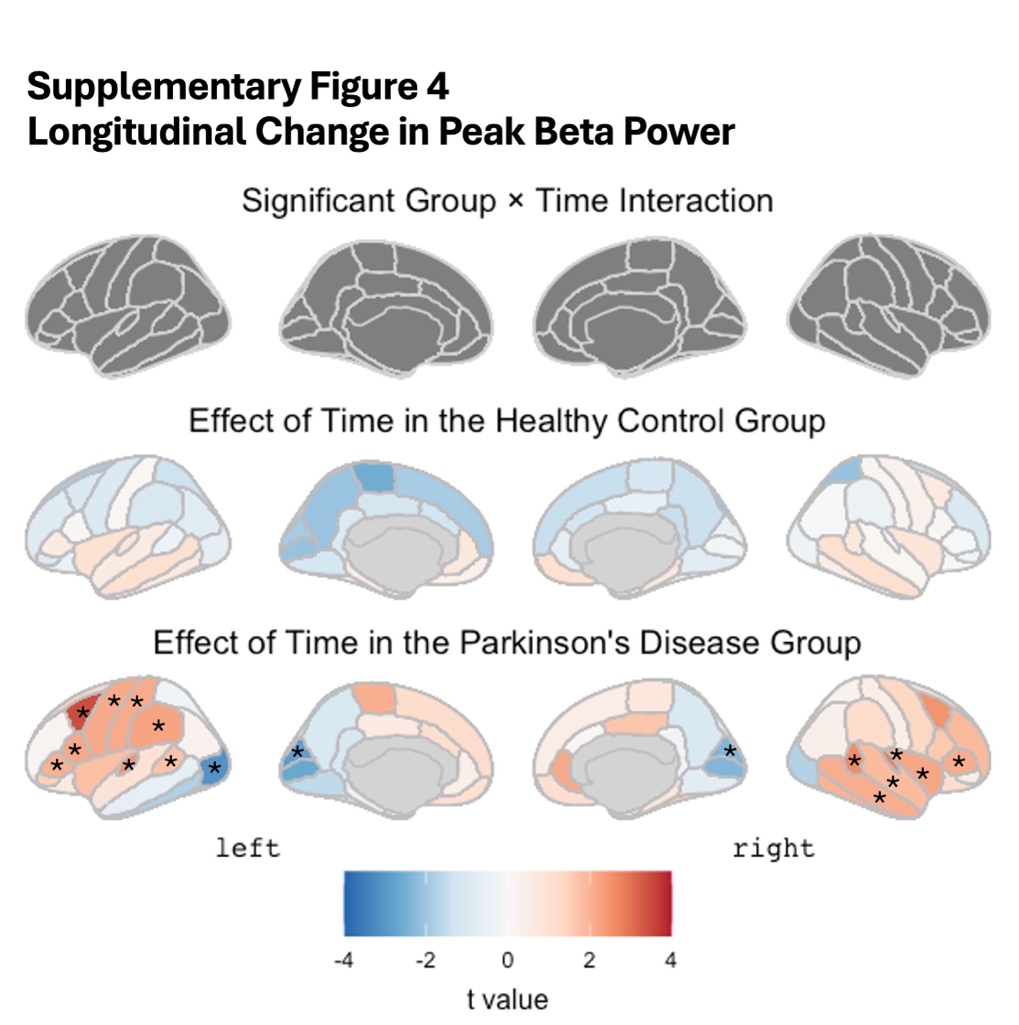

**Supplementary Figure 4: Longitudinal change of peak power in the beta frequency range. Upper row:** No evidence for significant group x time interaction when correcting for multiple testing with p_FDR_ < 0.05 across 68 brain regions. **Lower rows:** Main effects of time per group for illustration of the overall trends over time. * indicates regions with a significant effect of time with an FDR-corrected p-value < 0.05. Colouring represents the respective t values.

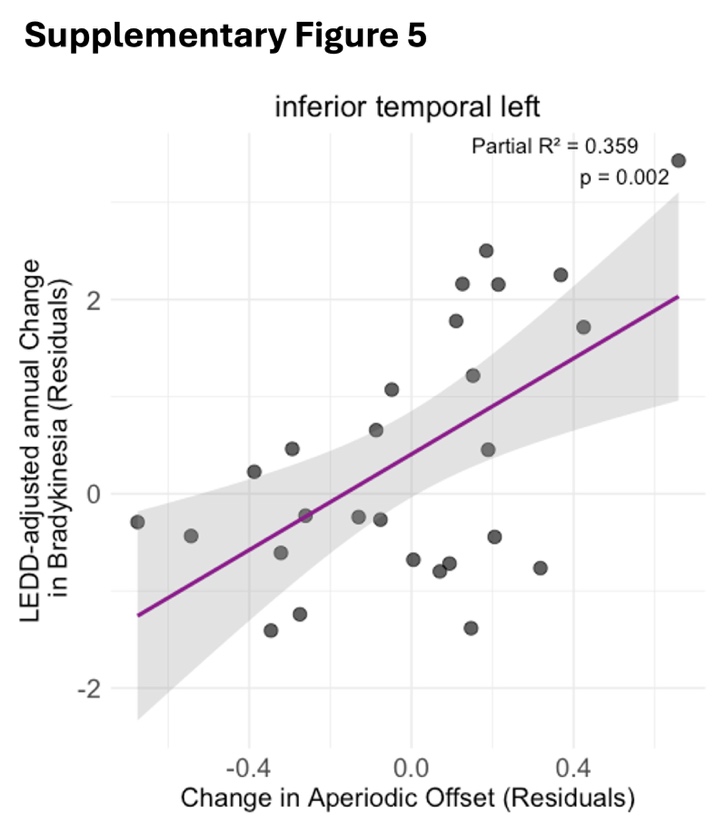

**Supplementary Figure 5: Relationship between longitudinal change of the aperiodic offset and progression of bradykinesia.** Residuals derived from the ANCOVA model described in the main text. R² in the upper right corner is the partial R², indicating how much of the variance in clinical change is explained by adding change in the aperiodic exponent to the model. The grey area represents the 95%-confidence interval around the fitted regression line in pink.

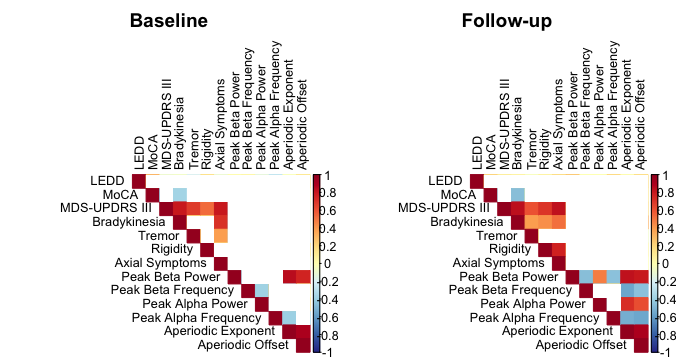

**Supplementary Figure 6: Correlation matrix for the relationships between clinical and neurophysiological variables at baseline (left) and follow-up.** Correlation coefficients are shown color-coded only for pairings with a significant correlation with uncorrected p < 0.05.

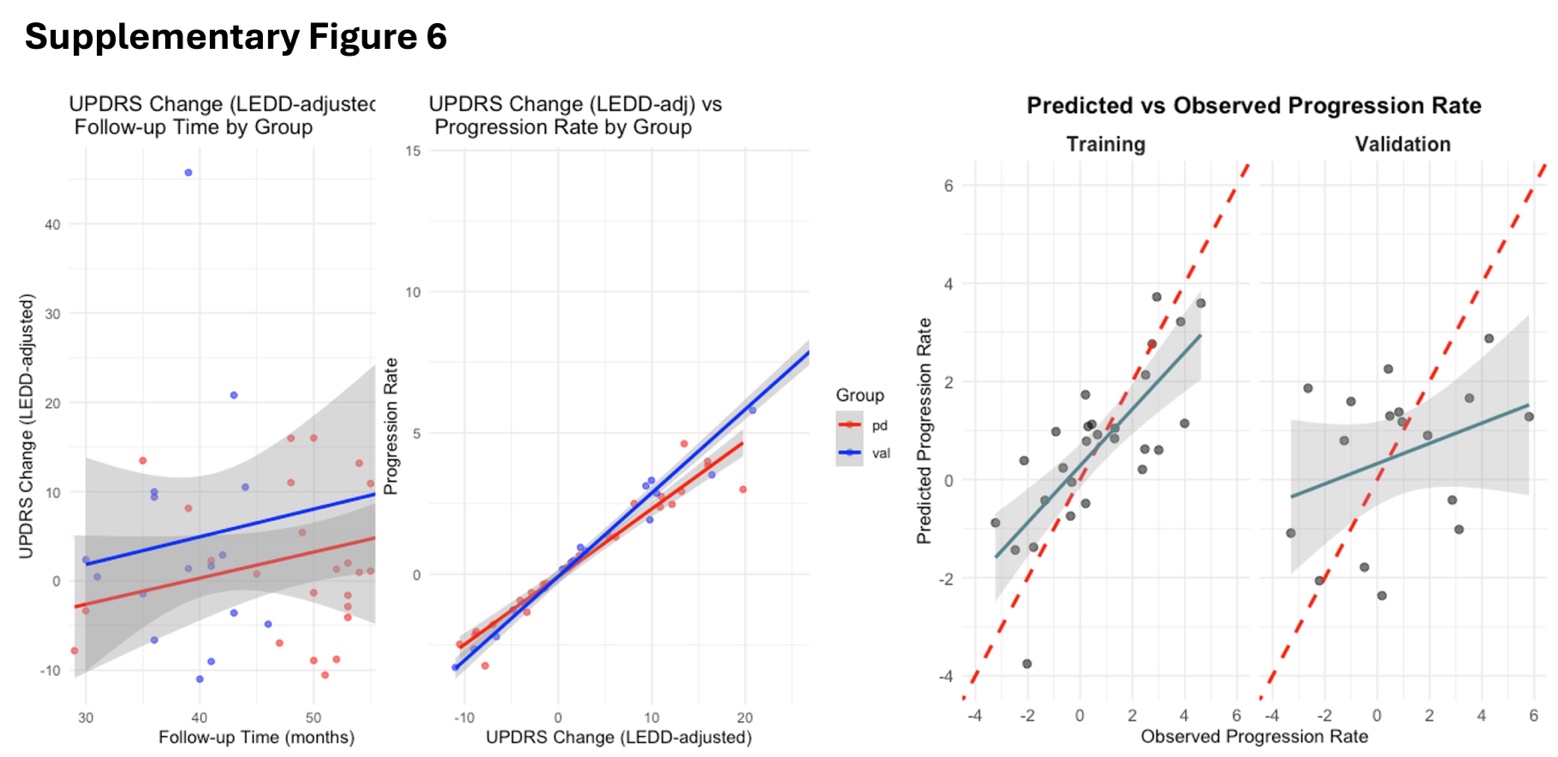

**Supplementary Figure 7: Left:** Relationship between time to follow-up and LEDD-adjusted annual change in MDS-UPDRS III score in the training (red) and validation cohort (blue). Note that the slopes run in parallel but with slightly higher motor symptom burden in the validation cohort. Second left: Correlation between the LEDD-adjusted annual change with the absolute change in MDS-UPDRS III scores for both groups. **Right:** Predicted versus actual observed progression rate in the training and validation cohorts.

**Supplementary Table 1:** Demographics and clinical characteristics of the validation cohort compared with the training cohort.

| **Characteristic** | **Training cohort**  (N = 27)*^1^* | **Validation cohort**  (N = 18)*^1^* | **t-statistic***^2^* | **P-value***^2^* |
| --- | --- | --- | --- | --- |
| **Age (years)** | 66.1 (10.2) | 65.4 (10.2) | t (43) = -0.24 | 0.8 |
| **Sex** | 12 (44%) | 8 (44%) | χ² (1) = 0 | >0.9 |
| **Disease Duration**  **at Follow-up (years)** | 9.1 (3.6) | 9.2 (2.0) | t (43) = 0.16 | 0.3 |
| **Time to Follow-up (months)** | 49.6 (9.6) | 40.6 (8.2) | t (43) = -3.27 | 0.002 |
| **MoCA Baseline** | 27.3 (2.3) | 25.5 (2.8) | t (42) = -2.38 | 0.031 |
| **MDS-UPDRS III Baseline** | 17.8 (9.5) | 19.1 (12.1) | t (43) = 0.38 | >0.9 |
| **MDS-UPDRS III Follow-up** | 19.3 (9.7) | 23.4 (12.8) | t (43) = 1.24 | 0.4 |
| **LEDD Baseline (mg/day)** | 469.6 (270.2) | 536.7 (226.4) | t (43) = 0.87 | 0.4 |
| **LEDD Follow-up (mg/day)** | 857.0 (293.3) | 740.6 (220.1) | t (43) = -1.44 | 0.2 |
| *^1^*Mean (SD); n (%) | | | | |
| *^2^*Within-group paired t-tests for validation cohort: MDS-UPDRS III: t (17) = -1.35, p = 0.195; LEDD: t (17) = -4.24, p = 0.001 | | | | |
